# Optimising a whole-genome sequencing workflow for One Health surveillance of Influenza A viruses across human, swine, and avian hosts in a diagnostic setting

**DOI:** 10.1101/2025.07.15.25331561

**Authors:** Klara M. Anker, Jesper S. Krog, Marta M. Ciucani, Ramona Trebbien

**Affiliations:** Department of Virus and Microbiological Special Diagnostics, Statens Serum Institut, Artillerivej 5, 2300 Copenhagen S, Denmark; Department of Health Technology, Technical University of Denmark, Ørsteds Plads, Building 345C, 2800 Kgs. Lyngby, Denmark

**Author notes:** **Correspondence**: Klara M. Anker.

**Keywords:** Influenza A virus, Whole genome sequencing, Genomic surveillance, One Health, One-tube RT-PCR, Next-generation Sequencing

## Abstract

Influenza A viruses (IAV) are a global health concern, infecting a wide range of hosts, including humans, birds, and pigs. Whole genome sequencing (WGS) is crucial for genomic surveillance under a One Health framework, providing insights into IAV evolution and transmission. We evaluated several laboratory workflows for next generation sequencing (NGS), aiming to optimise and unify the WGS process across IAV samples of human, swine, and avian origins.

Multiple combinations of RNA extraction methods, one-tube RT-PCR protocols and primer sets were systematically tested for their efficacy in generating high-quality PCR and NGS products across all viral segments. We assessed performance based on sequencing quality, including read counts, segment coverage and amplification bias across various host types, subtypes, and viral loads.

High-quality RNA extraction was critical for achieving reliable sequencing results, particularly in low viral load samples. Uniform amplification across all IAV genome segments was best achieved using an optimised RT-PCR protocol that reduced amplification bias for shorter fragments, including defective interfering particles (DIPs). Although variability in performance remained across host types and sample qualities, the refined workflow showed consistent improvements in sequencing outcomes, offering a promising foundation for a unified genomic surveillance. While further refinements may be needed for specific contexts, this study provides an improved, broadly applicable workflow that enhances genomic surveillance of influenza A viruses under a One Health framework, facilitating better understanding of viral evolution and transmission across species.

## Introduction

Influenza A virus (IAV) is a highly adaptable and continuously evolving pathogen. Its ability to cause seasonal and pandemic outbreaks in several hosts such as human, avian, and swine populations necessitates accurate and timely surveillance to inform effective control measures and antiviral strategies [1]. In this context, human and veterinary diagnostic laboratories play a pivotal role in the surveillance of IAVs circulating in both human and animal populations. Traditional methods apply real-time reverse transcriptase polymerase chain reaction (qRT-PCR) or Sanger sequencing for targeted molecular subtyping of Hemagglutinin and Neuraminidase genes [2,3]. However, the ability of IAV to undergo frequent reassortments and genetic drift calls for a more thorough and robust analysis of the entire viral genomes, and whole genome sequencing (WGS) methods are becoming more widely used in many surveillance laboratories [3]. The recent COVID-19 pandemic exemplified the important role of WGS in the monitoring of viral evolution, particularly in the detection of novel variants and mutations with influence on virulence, pathogenicity, and host range [4].

Given the zoonotic potential and close relationship between especially human and swine IAVs, the detailed insights provided by WGS have become an indispensable component of the surveillance and understanding of IAV transmissions between species in a One-Health perspective [1,3].

WGS of IAVs typically involves a multi segment RT-PCR strategy that allows for the simultaneous amplification of all eight IAV segments within a single one-step RT-PCR reaction, employing a universal primer set that targets the 5’ and 3’ ends conserved between each IAV segment [5,6]. Previous optimisation efforts of this one-tube RT-PCR workflow has focused on enhancing the initial RNA extraction [7,8] or comparing varying RT-PCR kits or primer combinations for better and more robust next generation sequencing (NGS) results [7,9–11]. However, these optimisation efforts have often been confined to specific IAV subtypes or host origins and on a limited number of samples. In a One-Health perspective, reference laboratories for both human and animal IAVs, need an efficient and reliable WGS workflow capable of delivering high-quality sequence data from all IAV subtypes and host origins. Thus, the main objective of this study was to align and optimise viral RNA extraction and one-tube RT-PCR protocols for consistent and successful WGS of IAV samples spanning human, swine, and avian origins, inclusive of various subtypes, sample types, and viral loads encountered in a diagnostic setting. As part of our continuous effort to enhance sequencing outputs, we tested different strategies for optimising both the extraction and one-tube RT-PCR of IAV segments before Nextera XT library preparation and sequencing on the Illumina platform.

## Methods

### Selection of samples

The samples used in this study of human, swine, and avian origin were all received as part of both active and passive influenza surveillance programs and previously identified as positive for influenza A virus. The samples were collected between 2020-2024 and had been stored at -80°C. A list of all the samples, including sample types, IAV subtypes, and Ct values is shown in **Table S1**. For samples extracted using both extraction method 1 (EX1) and extraction method 2 (EX2), the original sample material was diluted in PBS to have enough material for both extractions.

### Nucleic acid extraction

Total nucleic acid or RNA from the viral samples were extracted using either the MagNA Pure (EX1) or the QIAcube (EX2) protocols. For EX1, nucleic acid was extracted using a MagNA Pure 96 extraction robot (Roche) using the MagNA Pure 96 DNA and Viral NA Small Volume Kit and the Viral NA plasma ext lys SV 4.0 protocol following the manufacturer’s instructions (450 µl input and 100 µl elution volume).

For EX2, total RNA was extracted with RNeasy Mini Kit (QIAGEN, Denmark) automated on the QIAcube robot (QIAGEN) according to the instructions provided by the manufacturer. The samples were prepared for extraction as follows: 200 µl sample material was mixed with 400 µl RLT-buffer (included in the RNeasy kit) containing β–mercaptoethanol.

### Real-time quantitative reverse transcription polymerase chain reaction (RT-qPCR)

As an estimate of the viral load, extracted RNA from each sample was analysed in an MP-specific RT-qPCR assay [12]. Samples with cycle threshold (Ct) values >36 were excluded from further analysis and comparisons to reduce bias from low-quality samples on sequencing outcomes.

### One-tube Reverse Transcription Polymerase Chain Reaction (RT-PCR)

We evaluated a total of 5 different one-tube RT-PCR procedures previously described in literature. Some of these were also tested in combination with different primer sets. After preliminary assessments, we narrowed our focus to two methods, OT1 and OT2, detailed in **Table 1**, for which the majority of the WGS analyses were performed. The other tested methods, OT3-OT5, are described in **Table S3**.

**Table 1:** Description of the one-tube RT-PCR methods (OT1 and OT2) that we compared in this study. For additional tested methods, see Supplementary Table S3.

|  | **OT1**<br>(modified from [6]) | **OT2**<br>(modified from [7]) |
| --- | --- | --- |
| **RNA input** | 4 µl | 3 µl |
| **One-Step RT-PCR**<br>**Polymerase kit (enzyme**<br>**volume)** | SuperScript™ III One-Step RT-PCR System<br>with Platinum Taq High Fidelity DNA<br>Polymerase (Invitrogen™) (0.8 µl) | qScript® XLT One-Step RT-PCR Kit<br>(QuantaBio) (1 µl) |
| **Reaction Volume** | 40 µl | 25 µl |
| **Primers (concentration)**<br>(see Supplementary Table 2<br>for primer sequences) | MBTuni-12(R) (200 nM)<br>MBTuni-13 (200 nM)<br>or<br>MBTuni-12(A) (100 nM)<br>MBTuni-12(G) (100 nM)<br>MBTuni-13 (200 nM)<br>or<br>MBTuni-12(A) (80 nM)<br>MBTuni-12(G) (120 nM)<br>MBTuni-13 (200 nM)<br>or<br>Uni-12/Inf-1 (80 nM) | MBTuni-12(A) (400 nM)<br>MBTuni-12(G) (400 nM)<br>MBTuni-13 (800 nM)<br>or<br>Uni-12/Inf-1 (320 nM)<br>Uni-12/Inf-3 (480 nM)<br>Uni-13/Inf-1 (800 nM) |
|  | Uni-12/Inf-3 (120 nM) |  |
|  | Uni-13/Inf-1 (200 nM) |  |
| **Other reagents** |  | 1 mM MgCl<sub>2</sub> |
| **Thermocycler program** | 42°C for 60 min; 94°C for 2 min | 48°C for 20 min; 94°C for 3 min |
|  | [94°C for 30 sec; 45°C for 30 sec; 68°C for 3 min] x 5 | [94°C for 30 sec; 45°C for 30 sec; 68°C for 3 min] x 5 |
|  | [94°C for 30 sec; 57°C for 30 sec; 68°C for 3 min] x 31 | [94°C for 30 sec; 58°C for 30 sec; 68°C for 3 min] x 31 |
|  | 68°C for 7 min | 68°C for 10 min |

Based on previous experience and for higher consistency between the methods, we changed the primer set originally used in OT1 from a MBTuni-12(R)/MBTuni-13 set to one containing MBTuni-12(A), MBTuni-12(G) and MBTuni-13 primers in a 1:1:2 ratio (from now on called MBT primers) (**Table S2)**. The UNI primers (Uni-12/Inf1, Uni-12/Inf-3 and Uni-13/Inf-1) were used together with the qScript RT-PCR kit in the study by McGinnis et al. [7], so we tested these primers in the same relative concentration (0.8:1.2:2 ratio) of the primers as reported in the study (**Table S2)**. We introduced slight modifications to the original method proposed by McGinnis et al. [7], including a 4x increased primer concentration in what we describe as OT2. A second ratio of the MBT primer set, corresponding to the 0.8:1.2:2 ratio of the UNI primers was also tested on a subset of samples and referred to as MBT2.

Following the one-tube RT-PCRs, PCR products were visualised on Agarose gels and irrespective of the visual results, the remaining PCR products were purified using the GFX^TM^ PCR DNA and Gel Band Purification kit (Cytiva) using the manufacturer’s instructions. The final elution volume ranged from 10-50 µl; a smaller elution volume was used for samples where gels appeared blank or had smeared or indistinct bands, aiming for a more concentrated sample.

### Testing strategy

We initially compared two methods for extracting IAV RNA, EX1 (magnetic beads) and EX2 (Silica membrane), as these are the methods and kits routinely used in our laboratories. We extracted RNA from three human, four swine, and three avian origin samples of varying viral load based on previous RT-qPCR analyses (**Table S1,** AV, SW and HU w, m and s samples). We classified these into either weak (w), medium (m) or strong (s) samples based on the general span of Ct values observed for each host category in our surveillance. Following extraction, all samples were amplified using a modified version of the protocol described by Zhou et al. [6] (OT1) with MBT12(R)/MBT13 primers (MBTR, **Table S2**) and sequenced on the Illumina MiSeq platform.

We next tested the combination of the two extraction methods with different one-tube RT-PCR protocols and primers. Initial tests had indicated better performance of especially two RT-PCR methods: OT1 and OT2 (**Table 1**) when comparing PCR-fragments on agarose gels (**Figure S1**) or NGS results (**Figure S2**). Therefore, these two methods were used for further analyses in combination with two different sets of primers (MBT and UNI, see **Table S2**). We tested these methods on samples extracted using either EX1 or EX2; initially, 13 samples from each host were extracted using EX1 and 10 extra samples from each host (23 total from each host) were extracted using EX2, however, some of these samples were excluded from the final comparisons due to high Ct values (here, everything with a Ct >36) (see **Table S1**).

### DNA library preparation and Next Generation Sequencing

We quantified the purified PCR products using the Quant-iT™ dsDNA Broad-Range Assay Kit (Invitrogen™) and diluted to 0.2 ng/µl. Sequencing libraries were prepared using the Nextera® XT DNA Library Preparation kit (Illumina®), and the libraries were then quantified using the Quant-iT™ dsDNA High-Sensitivity Assay Kit and size-estimated using the TapeStation instrument (Agilent). Libraries were then normalised to 2nM, pooled and denatured, and the final libraries were diluted to 8 or 10 pM and spiked with 1% PhiX before sequencing on the Illumina MiSeq platform using the MiSeq V2 500 cycles kit.

### WGS analysis

The raw NGS reads were processed, filtered and assembled using the IRMA v.1.0.2 (Iterative Refinement Meta-Assembler) pipeline [13] to reconstruct consensus sequences of the samples and generate coverage tables and summary statistics. Reads with a quality score below 30 and a length below 125 bp were discarded, and a minimum of 50x coverage was used to call the consensus sequence. In case of coverage below the minimum threshold a missing site “N” was called.

The number of missing sites and the average read quality were used to determine the number of segments that would be considered successful (“passed”) during sequencing. When more than 10% of sites were missing or the average read quality was below 30, the segment would fail.

Tables containing information on the total reads counts, number of influenza-specific reads and the number of segment-specific reads were used as comparison between the different laboratory protocols tested in the study, and these were analysed and plotted in R [14] using the Tidyverse and ggplot2 packages [15,16].

## Results

### Comparison of two RNA extraction methods for influenza A virus sequencing

Initial tests conducted on a small set of samples from human (n=3), swine (n=4), and avian (n=3) (**Table S1**) hosts demonstrated a better overall NGS performance from samples extracted using the EX2 method. Despite comparable total read counts between the two extraction methods, samples extracted with EX2 yielded a higher percentage of influenza-specific reads and produced higher quality consensus sequences, resulting in more segments passing quality thresholds (**Figure 1a-c**). For samples with high viral loads (AV-s, SW-s, HU-s), both extraction methods provided sufficient read depth across most segments. However, for several of the medium and weak samples, only the EX2 method consistently generated sufficient reads to cover most viral segments. In contrast, the EX1 method often failed to produce adequate reads to cover any or more than a single segment for both the AV-w, HU-w and HU-m samples (**Figure 1d**).

**Figure 1:**
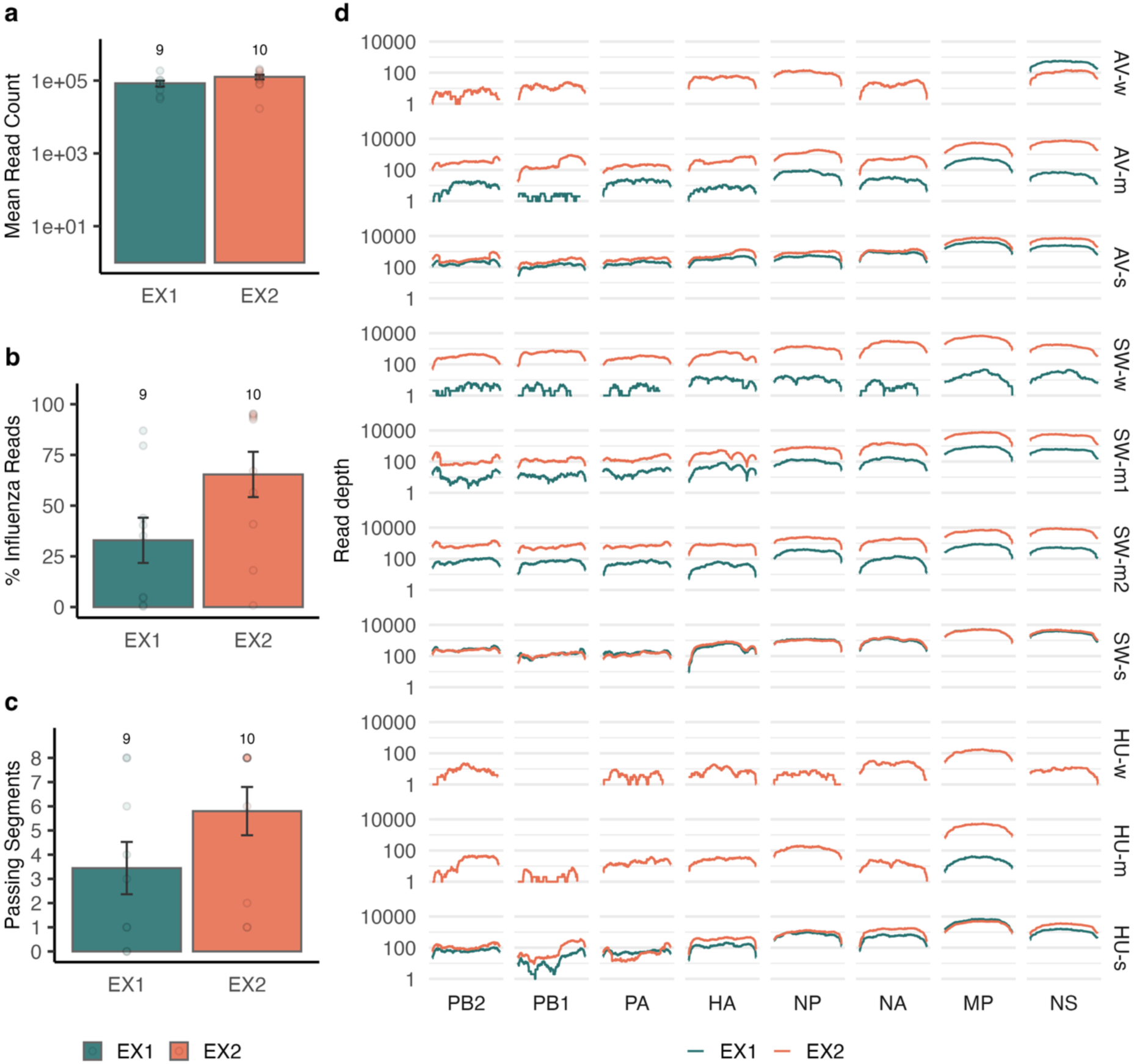
NGS read performance of IAV samples extracted with MagNA Pure (EX1) and QIAcube (EX2). **a.** Average total read counts across all samples for each extraction method. The y-axis is presented on a log10 scale for better visibility. **b.** Average percentage of Influenza-specific reads across all samples for each extraction method. **c.** Average number of IAV segments with complete, high quality consensus sequences (average read quality >30 and <10% missing bases) across all samples for each extraction method. Error bars show standard error of the mean and numbers above each bar show the number of samples contributing to the mean and standard error. One sample (HU-w) produced no usable reads following the EX1 extraction. **d.** Read depth across the lengths of the 8 IAV segments for each of the tested samples. The y-axis is presented on a log10 scale for better visibility (AV: avian origin, SW: swine origin, HU: human origin, w: weak sample (high Ct), m: medium sample, s: strong sample (low Ct). For excact ct values, see **Table S1**.

While this implied a clear influence of RNA quality on the final NGS outcomes, we next expanded our analysis to a larger sample set, evaluating combinations of extraction methods and one-tube RT-PCR protocols with different primer sets to identify the most robust approach for WGS across diverse sample types.

### Evaluation of RNA extraction methods and one-tube RT-PCR protocols

We assessed the performance of five one-tube RT-PCR protocols in total and narrowed these down to two (OT1 and OT2), that exhibited superior performance (**Figure S1, S2**). These two protocols were further expanded on by testing them with two different sets of primers (MBT and UNI) across samples of human, swine, and avian origins, following viral RNA extraction using either EX1 or EX2. While total read counts were very similar across all method combinations (**Figure 2a**), slight differences were observed in the proportion of influenza-specific reads depending on the extraction, RT-PCR method, and sample origin. Specifically, EX2 extraction generally resulted in higher influenza-specific reads counts for swine and avian samples, while EX1 performed slightly better for the human samples (**Figure 2b**). Among the RT-PCR and primer combinations, OT1-MBT tended to yield more influenza-specific reads, closely followed by the other methods, particularly OT1-UNI.

**Figure 2:**
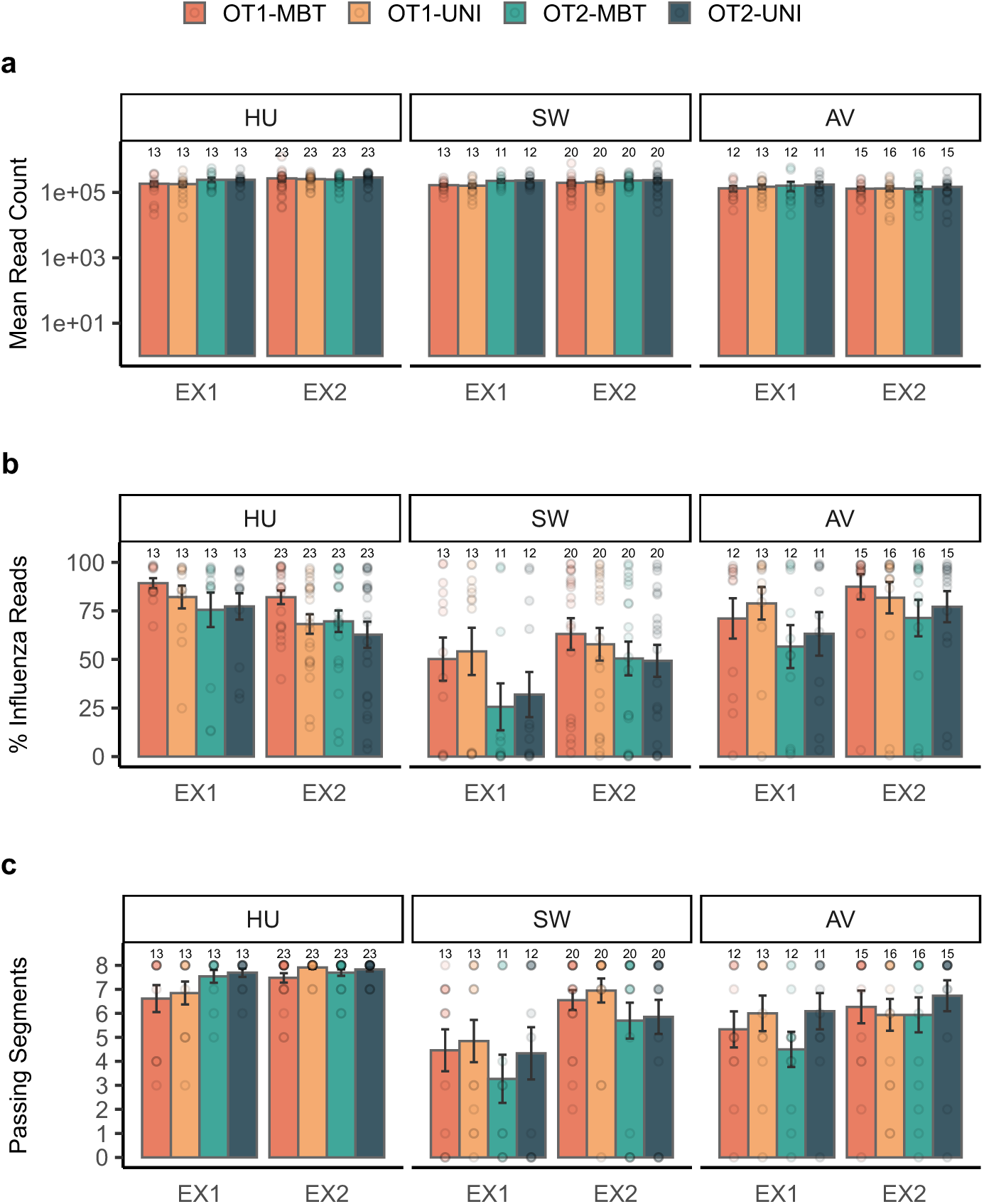
NGS read performance of IAV samples of human (HU), swine (SW), or avian (AV) origin extracted using EX1 or EX2 and amplified using two different one-tube RT-PCR methods and two different sets of primers. **a.** Average total read counts across all samples for each extraction method. The y-axis is presented on a log10 scale for better visibility. **b.** Average percentage of influenza-specific reads across all samples for each extraction method. **c.** Average number of IAV segments with complete, high quality consensus sequences passing quality filters (average read quality >30 and <10% missing bases) across all samples for each extraction method. Points represent each sample, and the total number of samples used to calculate the mean values is indicated above each bar. Error bars represent the standard error of the mean.

The number of segments passing quality filters was generally higher for EX2 than for EX1 extracted samples across all host origins (**Figure 2c**). Nearly every segment passed for the human-origin samples, and there was a clear improvement in passing segments in swine-origin samples following EX2 compared to EX1 extraction. Although the RT-PCR protocol had less pronounced effect on the total number of passing segments, OT2-UNI showed a slightly higher average number of passing segments overall, except in the swine-origin samples.

### Read distribution and quality across IAV segments

The distribution of influenza-specific reads across the eight IAV segments varied depending on the RT-PCR method and primer set used. Samples of all host origins processed with OT1-MBT exhibited a skewed read distribution, with a considerably higher proportion of reads mapping to the shorter MP and NS segments, while fewer reads were assigned to the longer polymerase segments (PB2, PB1, PA) and HA (**Figure 3a**). In contrast, samples processed with OT2-UNI showed a more balanced read distribution across all segments, though minor fluctuations remained. Notably, the OT1 or OT2 methods combined with the UNI primers appeared to improve the proportion of reads mapping to the polymerase segments, suggesting an advantage in amplifying the longer viral segments. Similar overall distribution pattern of reads between segments for each method was observed for samples extracted with EX1 and EX2, indicating that this distribution was more affected by the one-tube RT-PCR and primers and not the extracted RNA.

**Figure 3:**
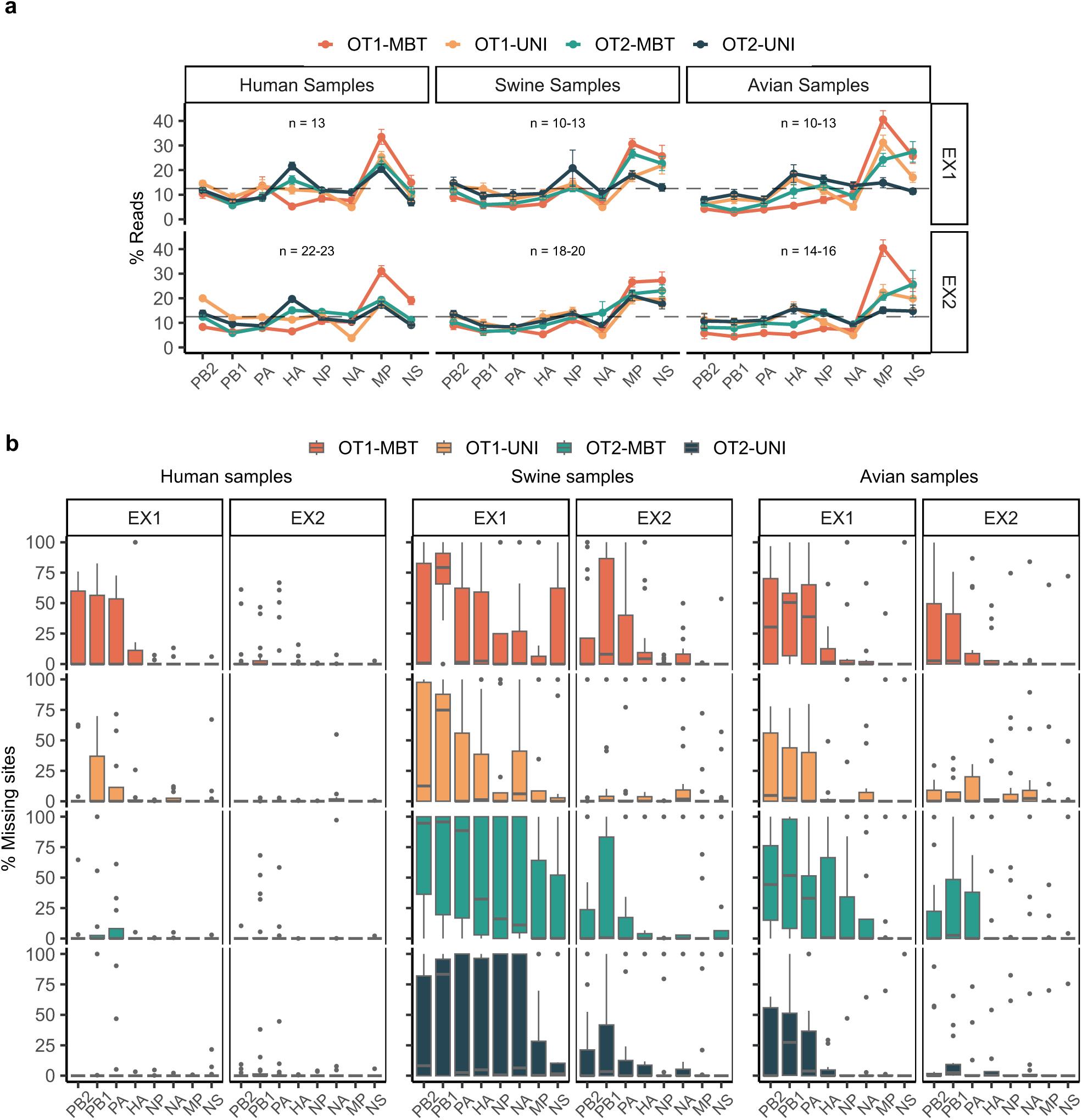
Read distribution and quality of consensus segments following NGS of IAV samples of human, swine, or avian origin extracted using EX1 or EX2 and amplified with two different one-tube RT-PCR methods and two different sets of primers **a.** Average percentage of influenza-specific reads in a sample that mapped to each IAV segment. Coloured lines connect the average read percentages per segment for each RT-PCR method for better visualization. The number of samples used to calculate the mean values are indicated above each plot, sometimes spanning more values when some segments were missing from one or more samples or methods. Error bars represent the standard error of the mean, and the dotted line indicates the 12.5% resulting from an equal distribution of reads to each of the 8 segments. **b.** Boxplot showing the percentage of missing sites in consensus sequences of each IAV segment. The number of samples included is not shown on the boxplots, but is similar to the numbers in **a**; Human samples EX1: n = 13, EX2: n = 22-23; Swine samples EX1: n = 10-13, EX2: n = 18-20; Avian samples EX1: n = 10-13, EX2: n = 14-16.

Read quality across segments also showed distinct patterns based on extraction method, RT-PCR protocol and sample origin. While human samples generally yielded high-quality consensus sequences with almost no missing bases (N’s) regardless of the method used (**Figure 3b**), swine and avian samples often generated lower-quality sequences, particularly for the polymerase segments. There was a clear tendency of more missing sites for samples extracted using EX1 compared to EX2, especially for swine and avian samples, and for human OT1-MBT or OT1-UNI processed samples. Across all sample types, EX2 extraction in combination with the OT2-UNI or OT1-UNI protocol resulted in fewer missing sites across all segments.

We observed a more even read distribution and fewer missing sites, particularly in the polymerase segments, when using the UNI primers with both the OT1 and OT2 protocols. This led us to hypothesise that the uneven ratio of forward primers used in the UNI primer set might favour amplification of the polymerase segments, which are known to frequently contain a cytosine at the 4^th^ position of the conserved 3’ end, compared to the uridine present in other segments [17,18]. To test the influence of the primer ratio, we adjusted the MBT primer set to match the relative concentrations of the UNI primers in OT1 (OT1-MBT2) and tested this on a subset of samples. However, this adjustment did not yield the same improvements for the polymerase segments (**Figure S3**). Instead, OT1-MBT2 resulted in fewer segments passing quality thresholds for both human and avian samples (**Figure S3a**), inconsistent read distributions across segments with an overrepresentation of the MP segment (**Figure S3b**), and more missing sites, particularly in human and avian samples (**Figure S3c**).

### Read coverage patterns across segments

Lastly, we assessed the read coverage patterns across IAV segments to better understand how the different extraction methods and RT-PCR protocols influenced genome completeness. Generally, samples extracted using EX2 exhibited more consistent and higher average read depth across most segments compared to those extracted with EX1, regardless of the RT-PCR protocol used (**Figure 4**). Among the RT-PCR methods, OT2-UNI consistently provided higher and more even coverage across most segments (except the MP and NS segments) compared to the other protocols, reflecting the more balanced read distribution previously observed (**Figure 3a**).

**Figure 4:**
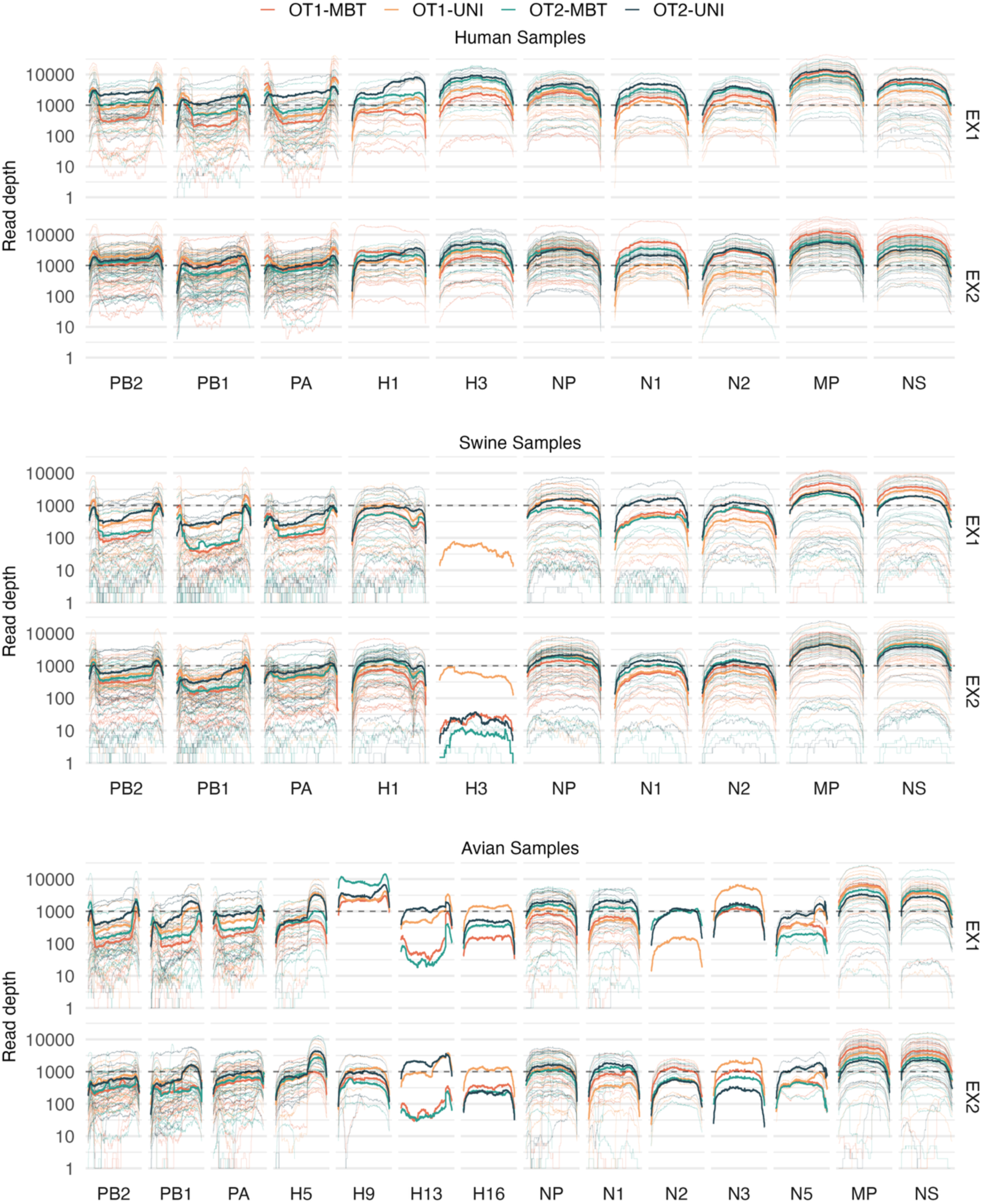
Coverage plots showing the number of reads at each position of each segment for all samples extracted using EX1 or EX2 and amplified with two different one-tube RT-PCR methods and two different sets of primers. Thin lines each represent the coverage of one sample for each segment, while the thick lines represent the average coverage observed for all samples treated with a specific one-tube RT-PCR method. The y-axis is presented on a log10 scale, and a dotted line shows a read depth of 1000 reads for better comparison between segments.

A recurring pattern observed across all methods, particularly in samples processed with OT1-MBT, was the presence of high coverage peaks at the 5’ and 3’ ends of the polymerase segments (PB2, PB1, and PA), resulting in noticeable drops in coverage in their central regions. This tendency was especially pronounced in swine-origin samples and seemed to diminish in samples extracted with EX2, where the coverage was more uniform across these segments. Interestingly, samples processed with OT2-UNI generally showed a much more even coverage profile, with only minor peaks at the ends of the polymerase segments, irrespective of the extraction method.

Another notable observation was the occurrence of localised coverage peaks in the middle of specific segments in some avian-origin samples. This was most pronounced in the PB1 and H5 segments, particularly in samples treated with OT1-UNI and OT2-UNI, as well as in some OT2-MBT treated samples for H5. These peaks were less evident or absent in samples treated with OT1-MBT. Similar patterns were seen in human-origin H1 segments and swine-origin PB1 segments, with more pronounced peaks in samples extracted using EX1 than EX2.

Samples processed with OT1-MBT2 did not show these mid-segment peaks but exhibited a similar overall coverage pattern as samples treated with OT1-MBT, including the central coverage drops in the polymerase segments (**Figure S4**). However, OT1-MB2 yielded lower overall coverage across segments and host types than any of the other RT-PCR methods tested.

## Discussion

Our analysis demonstrates the major impact of RNA extraction methods and one-tube RT-PCR protocols on the quality and completeness of IAV WGS from samples of human, swine, and avian origin. In line with concerns raised in previous studies [11], the variability in WGS results across IAV samples and the absence of standardised procedures motivated us to refine the one-tube RT-PCR workflow and establish an optimised methodology for consistent and robust surveillance across diverse host origins within the One-Health framework.

A key finding of our study was the considerable influence of extraction methods on WGS performance. Specifically, we observed an overall superior performance of the EX2 method compared to EX1, particularly for samples with low viral loads or poorer quality of original sample material, more frequently encountered in swine or avian surveillance samples. This suggests that the silica membrane-based approach of EX2 is more effective in minimising contaminants and enhancing RNA purity and integrity, compared to the magnetic particle-based approach in EX1, which could potentially lead to shearing of longer RNA fragments. Furthermore, the EX1 method, which extracts total nucleic acids, may result in higher proportions of inhibitory DNA in the final eluate, in contrast to EX2, which is tailored specifically for RNA extraction. Di et al. [8] previously assessed IAV extraction and subsequent PCR amplification and WGS of nasal wash samples from ferrets inoculated with avian IAVs. Their findings emphasised the importance of specific RNA extraction and DNase treatment in achieving high quality NGS data across all IAV segments and showed that elevated levels of background DNA directly impeded the PCR amplification of IAV segments, even when using IAV-specific primers. Although they favoured the MagNA Pure compact RNA isolation kit over the RNeasy Mini kit used in our EX2 method, no analogous RNA-specific extraction kit or protocol is available for the high-throughput MagNA Pure 96 system. Thus, there appears to be a trade-off between the high-throughput capability of EX1 and the higher RNA quality achievable with EX2, depending on the specific sample type, quality and research scope. Separate DNase treatment, as shown by Di et al. [8] to restore IAV RNA quality, is not feasible in laboratories that also routinely test and analyse DNA viruses, and although Trizol-like extractions are generally preferred for optimal RNA quality, they are not applicable to the high-throughput diagnostic setting for which this workflow was optimised.

Although the tested RT-PCR methods produced comparable results in terms of the percentage of influenza-specific reads and the average number of high-quality recovered segments, notable differences were observed in read distribution and segment coverage. The OT2-UNI method consistently yielded high and uniform coverage across all eight segments, outperforming the widely used OT1-MBT method, which showed skewed read distributions, favouring shorter RNA fragments. This skewed distribution was reflected in the high coverage of MP and NS segments and the significant coverage drops in the central regions of the polymerase segments. These drops are likely caused by preferential amplification of defective interfering particles (DIPs), which contain large internal deletions and are known to especially arise from the longer polymerase segments [19–22]. DIPs have been shown to interfere with wild-type virus replication and trigger host immune responses, which could have profound implications for the virus-host interactions and disease dynamics [23,24]. Therefore, the high prevalence of DIPs, especially observed in our swine-origin samples, underscores a potentially distinct dynamic of the IAV infections across host or sample types that deserves further investigation.

The OT2-UNI method notably reduced the amplification bias towards DIPs and provided more uniform coverage across all segments, crucial for generating high-quality consensus sequences of all segments. However, occasional coverage peaks in the HA and PB1 segments, potentially due to off-target primer binding, were observed when using the UNI primers. This effect was most pronounced in avian-origin H5 segments, and to a lesser extent, in human-origin H1 and swine-origin PB1 segments. Despite these peaks, the overall coverage across segments remained comparable or higher than that achieved with other methods.

Our study further highlights the importance of representing sequence variations in the universal IAV primer set. The OT3 method, which employed a single forward MBT-uni-12(A) primer, struggled to amplify and provide sequencing reads for the polymerase segments, particularly PB1 and PA. This aligns with the documented variability of the 4th position of the conserved 12-nucleotide 3’ end of IAV segments, where the PB2, PB1 and PA segments more frequently contain a cytosine rather than a uridine [17,18]. Thus, including primers that account for this variability appears essential for successful amplification of full-length polymerase segments across all host types. Although we hypothesised that the improved polymerase segment coverage with UNI primers might be due to the imbalanced ratio of forward primers (position 4 A and G variants), adjusting the primer ratio accordingly for the MBT primers with the OT1 protocol did not yield similar improvements in our study.

Our evaluation was anchored in the protocols and procedures routinely employed by our reference laboratory for human and animal IAVs. Nevertheless, alternative technologies may address some of the challenges we identified. Notably, Nanopore technology, with its ability to generate longer reads and provide real-time data output, emerges as a valuable tool, especially when rapid sequencing results are needed [25–27]. Furthermore, Nanopore’s capability for direct RNA sequencing offers the potential to capture previously overlooked regions of the influenza genome, including the non-coding ends 3’ and 5’ ends, which are fundamental for our current PCR strategies [28–30]. Insights into potential variations in these regions could enhance our understanding of IAV infection dynamics, paving the way for more comprehensive surveillance and research in the future.

While our study demonstrates that the EX2 extraction method combined with the OT2-UNI protocol yields superior results across host types, further refinements and testing may enhance performance for specific sample types or high-throughput settings. Nonetheless, this approach offers a promising foundation for a unified workflow in IAV genomic surveillance

### Ethical statement

No ethical approval was required for this study. All samples were anonymised prior to analysis.

## Use of artificial intelligence tools

AI tools, specifically ChatGPT by OpenAI, were used to assist with text refinements, including improving clarity, conciseness, and overall flow of the manuscript. All scientific content, analysis, and conclusions were developed by the authors.

## Data availability

The scripts and summary data files, including read counts, segment coverage, and sequencing performance metrics, used in the analysis are available at https://github.com/KMAnker/One_Health_OneTube.

IAV consensus sequences from our surveillance samples are regularly uploaded to public databases like GISAID and GenBank, which also includes samples used in this analysis.

## Acknowledgements

We would like to acknowledge the laboratory technicians Carina Bøegh Folsing, Isabel Bro Høy, Jesper Rønn, Mai-Britt Jørgensen, Malika Winsløw Kolbak, Michala Cecilie Nowak and Sari Mia Dose from the National Influenza Centre at Statens Serum Institut for technical assistance in the laboratory.

## Authors’ contributions

**Klara M. Anker**: Conceptualization, Methodology, Software, Formal analysis, Investigation Visualization, Writing – original draft. **Jesper S. Krog**: Conceptualization, Methodology, Data curation, Project administration, Writing – review and editing. **Marta M. Ciucani**: Software, Formal analysis, Writing – review and editing. **Ramona Trebbien**: Conceptualization, Supervision, Funding acquisition, Writing – review and editing.

## References

[1] Harrington WN, Kackos CM, Webby RJ. The evolution and future of influenza pandemic preparedness. Exp Mol Med 2021;53:737–49. 10.1038/s12276-021-00603-0.

[2] Van Poelvoorde LAE, Saelens X, Thomas I, Roosens NH. Next-Generation Sequencing: An Eye-Opener for the Surveillance of Antiviral Resistance in Influenza. Trends Biotechnol 2020;38:360–7. 10.1016/j.tibtech.2019.09.009.

[3] Daniels RS, McCauley JW. The health of influenza surveillance and pandemic preparedness in the wake of the COVID-19 pandemic. J Gen Virol 2023;104:001822. 10.1099/jgv.0.001822.

[4] Oude Munnink BB, Worp N, Nieuwenhuijse DF, Sikkema RS, Haagmans B, Fouchier RAM, et al. The next phase of SARS-CoV-2 surveillance: real-time molecular epidemiology. Nat Med 2021;27:1518–24. 10.1038/s41591-021-01472-w.

[5] Hoffmann E, Stech J, Guan Y, Webster RG, Perez DR. Universal primer set for the full-length amplification of all influenza A viruses. Arch Virol 2001;146:2275–89. 10.1007/s007050170002.

[6] Zhou B, Donnelly ME, Scholes DT, St. George K, Hatta M, Kawaoka Y, et al. Single-Reaction Genomic Amplification Accelerates Sequencing and Vaccine Production for Classical and Swine Origin Human Influenza A Viruses. J Virol 2009;83:10309–13. 10.1128/JVI.01109-09.

[7] McGinnis J, Laplante J, Shudt M, George KS. Next generation sequencing for whole genome analysis and surveillance of influenza A viruses. Journal of Clinical Virology 2016;79:44–50. 10.1016/j.jcv.2016.03.005.

[8] Di H, Thor SW, Trujillo AA, Stark TJ, Marinova-Petkova A, Jones J, et al. Comparison of nucleic acid extraction methods for next-generation sequencing of avian influenza A virus from ferret respiratory samples. J Virol Methods 2019;270:95–105. 10.1016/j.jviromet.2019.04.014.

[9] Wang J. MinION nanopore sequencing of an influenza genome. Front Microbiol 2015;6:151109. 10.3389/fmicb.2015.00766.

[10] Lee HK, Lee CK, Tang JW-T, Loh TP, Koay ES-C. Contamination-controlled high-throughput whole genome sequencing for influenza A viruses using the MiSeq sequencer. Sci Rep 2016;6:33318. 10.1038/srep33318.

[11] Wüthrich D, Lang D, Müller NF, Neher RA, Stadler T, Egli A. Evaluation of two workflows for whole genome sequencing-based typing of influenza A viruses. J Virol Methods 2019;266:30–3. 10.1016/j.jviromet.2019.01.009.

[12] Nagy, A., Černíková, L., Kunteová, K., Dirbáková, Z., Thomas, S. S., Slomka, M. J., et al. A universal RT-qPCR assay for “One Health” detection of influenza A viruses. PloS one 2021;16(1):e0244669. 10.1371/journal.pone.0244669.

[13] Shepard SS, Meno S, Bahl J, Wilson MM, Barnes J, Neuhaus E. Viral deep sequencing needs an adaptive approach: IRMA, the iterative refinement meta-assembler. BMC Genomics 2016;17:708. 10.1186/s12864-016-3030-6.

[14] R Core Team. R: A Language and Environment for Statistical Computing. R Foundation for Statistical Computing, Vienna, Austria 2022.

[15] Wickham H, Averick M, Bryan J, Chang W, McGowan L, François R, et al. Welcome to the Tidyverse. J Open Source Softw 2019;4:1686. 10.21105/joss.01686.

[16] Wickham H. ggplot2. Cham: Springer International Publishing; 2016. 10.1007/978-3-319-24277-4.

[17] Seong BL, Lee KH. The position 4 nucleotide at the 3’ end of the influenza virus neuraminidase vRNA is involved in temporal regulation of transcription and replication of neuraminidase RNAs and affects the repertoire of influenza virus surface antigens. Journal of General Virology 1998;79:1923–34. 10.1099/0022-1317-79-8-1923.

[18] Benkaroun J, Robertson G, Whitney H, Lang A. Analysis of the Variability in the Non-Coding Regions of Influenza A Viruses. Vet Sci 2018;5:76. 10.3390/vetsci5030076.

[19] Davis AR, Hiti AL, Nayak DP. Influenza defective interfering viral RNA is formed by internal deletion of genomic RNA. Proceedings of the National Academy of Sciences 1980;77:215–9. 10.1073/pnas.77.1.215.

[20] Saira K, Lin X, DePasse J V., Halpin R, Twaddle A, Stockwell T, et al. Sequence analysis of in vivo defective interfering-like RNA of influenza A H1N1 pandemic virus. J Virol 2013;87:8064–74. 10.1128/JVI.00240-13.

[21] Mendes M, Russell AB. Library-based analysis reveals segment and length dependent characteristics of defective influenza genomes. PLoS Pathog 2021;17:e1010125. 10.1371/journal.ppat.1010125.

[22] Alnaji FG, Holmes JR, Rendon G, Vera JC, Fields CJ, Martin BE, et al. Sequencing Framework for the Sensitive Detection and Precise Mapping of Defective Interfering Particle-Associated Deletions across Influenza A and B Viruses. J Virol 2019;93. 10.1128/JVI.00354-19.

[23] Alnaji FG, Reiser WK, Rivera-Cardona J, Te Velthuis AJW, Brooke CB. Influenza A Virus Defective Viral Genomes Are Inefficiently Packaged into Virions Relative to Wild-Type Genomic RNAs. MBio 2021;12:e0295921. 10.1128/mBio.02959-21.

[24] Vignuzzi M, López CB. Defective viral genomes are key drivers of the virus–host interaction. Nat Microbiol 2019;4:1075–87. 10.1038/s41564-019-0465-y.

[25] King J, Harder T, Beer M, Pohlmann A. Rapid multiplex MinION nanopore sequencing workflow for Influenza A viruses. BMC Infect Dis 2020;20:648. 10.1186/s12879-020-05367-y.

[26] Crossley BM, Rejmanek D, Baroch J, Stanton JB, Young KT, Killian ML, et al. Nanopore sequencing as a rapid tool for identification and pathotyping of avian influenza A viruses. Journal of Veterinary Diagnostic Investigation 2021;33:253–60. 10.1177/1040638720984114.

[27] Vereecke N, Woźniak A, Pauwels M, Coppens S, Nauwynck H, Cybulski P, et al. Successful Whole Genome Nanopore Sequencing of Swine Influenza A Virus (swIAV) Directly from Oral Fluids Collected in Polish Pig Herds. Viruses 2023;15:435. 10.3390/v15020435.

[28] Keller MW, Rambo-Martin BL, Wilson MM, Ridenour CA, Shepard SS, Stark TJ, et al. Direct RNA Sequencing of the Coding Complete Influenza A Virus Genome. Sci Rep 2018;8:14408. 10.1038/s41598-018-32615-8.

[29] Lewandowski K, Xu Y, Pullan ST, Lumley SF, Foster D, Sanderson N, et al. Metagenomic Nanopore Sequencing of Influenza Virus Direct from Clinical Respiratory Samples. J Clin Microbiol 2019;58. 10.1128/JCM.00963-19.

[30] Perlas A, Reska T, Croville G, Tarrés-Freixas F, Guérin J-L, Majó N, et. al. Improvements in RNA and DNA nanopore sequencing allow for rapid genetic characterization of avian influenza. Virus Evolution, 2025;11:veaf010. 10.1093/ve/veaf010.

