## Supplemental Tables and Figures for "Optimising a whole-genome sequencing workflow for One Health surveillance of Influenza A viruses across human, swine, and avian hosts in a diagnostic setting"

### Supplementary tables and Figures

**Table S1**: List of all samples included in the analyses. Sample host origin, subtype, sample material and Ct values determined by real-time quantitative reverse transcription polymerase chain reaction (RT-qPCR) are shown along with indications of the extraction and One-tube RT-PCR methods the sample was tested in and whether the sample has been sequenced. Samples lacking NA subtype identification are indicated with an x (Nx). For most swine origin samples, the subtype is indicated as "pdm09" for H1N1pdm09 origin, "av" for "avian-like" swine H1N1 origin, or "sw" for swine H3N2 origin. Samples extracted using EX1 and EX2 have the respective Ct values listed as Ct EX1/Ct EX2, except when marked by *, which indicate Ct values from a previous RT-qPCR test following another extraction of the same sample. OT methods is as described in **Table 1** and **Supplementary Table 2** and is written either as full combinations of OT and primers or with a semicolon separating the OT and primers, indicating that all combinations of the listed OT methods and primers have been tested. Rows highlighted in grey indicate samples where sequences have been filtered out and don't contribute to comparisons of methods due to their high Ct values.

| Sample | Host | Subtype | Sample material | Ct value | EX methods | OT methods | Sequenced |
|---|---|---|---|---|---|---|---|
| AV-w | Avian | H5N1 | Cloacal swab | 38.8* | EX1, EX2 | OT1-MBTR | yes |
| AV-m | Avian | H5N1 | Cloacal swab | 31.1* | EX1, EX2 | OT1-MBTR | yes |
| AV-s | Avian | H5N1 | Brain swab | 21.1* | EX1, EX2 | OT1-MBTR | yes |
| SW-w | Swine | H1N2 | Pool of nasal swabs (x5) | 27.2* | EX1, EX2 | OT1-MBTR | yes |
| SW-m1 | Swine | H1N2 | Pool of nasal swabs (x5) | 23.8* | EX1, EX2 | OT1-MBTR | yes |
| SW-m2 | Swine | H1N1 | Nasal swab | 25.8* | EX1, EX2 | OT1-MBTR | yes |
| SW-s | Swine | H1N2 | Pool of nasal swabs (x?) | 22.0* | EX1, EX2 | OT1-MBTR | yes |
| HU-w | Human | H3N2 | Throat swab | 33.5* | EX1, EX2 | OT1-MBTR | yes |
| HU-m | Human | H3N2 | Throat swab | 24.8* | EX1, EX2 | OT1-MBTR | yes |
| HU-s | Human | H1N1pdm09 | Throat swab | 23.5* | EX1, EX2 | OT1-MBTR | yes |
| HU-1 | Human | H1N1pdm09 | Swab | 29.9/31.4 | EX1, EX2 | OT1, OT2; MBT, UNI | yes |
| HU-2 | Human | H3N2 | Swab | 30.1/33.2 | EX1, EX2 | OT1, OT2; MBT, UNI | yes |
| HU-3 | Human | H3N2 | Throat swab | 29.4/32.1 | EX1, EX2 | OT1, OT2; MBT, UNI | yes |
| HU-4 | Human | H1N1pdm09 | Throat swab | 35.6 | EX2 | OT1, OT2; MBT, UNI | yes |
| HU-5 | Human | H1N1pdm09 | Throat swab | 30.1 | EX2 | OT1, OT2; MBT, UNI | yes |
| HU-6 | Human | H1N1pdm09 | Throat swab | 34.5 | EX2 | OT1, OT2; MBT, UNI | yes |

| | | | | | | | |
|---|---|---|---|---|---|---|---|
| HU-7 | Human | H3N2 | Unknown | 30.9 | EX2 | OT1, OT2; MBT, UNI | yes |
| HU-8 | Human | H3N2 | Throat swab | 32.3 | EX2 | OT1, OT2; MBT, UNI | yes |
| HU-9 | Human | H3N2 | Unknown | 31.6 | EX2 | OT1, OT2; MBT, UNI | yes |
| HU-10 | Human | H3N2 | Throat swab | 32.1 | EX2 | OT1, OT2; MBT, UNI | yes |
| HU-11 | Human | H3N2 | Unknown | 33.3 | EX2 | OT1, OT2; MBT, UNI | yes |
| HU-12 | Human | H3N2 | Unknown | 32.4 | EX2 | OT1, OT2; MBT, UNI | yes |
| HU-13 | Human | H3N2 | Throat swab | 34.0 | EX2 | OT1, OT2; MBT, UNI | yes |
| HU-14 | Human | H3N2 | Unknown | 16.4 | EX1 | OT1-MBTR, OT2-UNI, OT3-MBTA, OT4-UNI | no |
| HU-15 | Human | H1N1pdm09 | Unknown | 17.2 | EX1 | OT1-MBTR, OT2-UNI, OT3-MBTA, OT4-UNI | no |
| HU-16 | Human | H1N1pdm09 | Throat swab | 28.1/29.8 | EX1, EX2 | OT1, OT2, OT5; MBT, UNI | no |
| HU-17 | Human | H1N1pdm09 | Throat swab | 31.0/32.8 | EX1, EX2 | OT1, OT2, OT5; MBT, UNI | no |
| HU-18 | Human | H1N1pdm09 | Throat swab | 30.9/31.3 | EX1, EX2 | OT1, OT2, OT5; MBT, UNI | no |
| HU-19 | Human | H1N1pdm09 | Unknown | 29.1/30.8 | EX1, EX2 | OT1, OT2, OT5; MBT, UNI | no |
| HU-20 | Human | H3N2 | Unknown | 27.6/30.4 | EX1, EX2 | OT1, OT2, OT5; MBT, UNI | no |
| HU-21 | Human | H3N2 | Unknown | 30.2/32.4 | EX1, EX2 | OT1, OT2, OT5; MBT, UNI | no |
| HU-22 | Human | H1N1pdm09 | Throat swab | 19.1 | EX1 | OT1-MBTR, OT2-UNI, OT3-MBTA | no |
| HU-23 | Human | H1N1pdm09 | Throat swab | 19.1 | EX1 | OT1-MBTR, OT2-UNI, OT3-MBTA | yes |
| HU-24 | Human | H1N1pdm09 | Unknown | 25.4 | EX1 | OT1-MBTR, OT2-UNI, OT3-MBTA | yes |
| HU-25 | Human | H3N2 | Swab | 29.1 | EX1 | OT1-MBTR, OT2-UNI, OT3-MBTA | yes |
| HU-26 | Human | H3N2 | Swab | 26.2 | EX1 | OT1-MBTR, OT2-UNI, OT3-MBTA | yes |
| HU-27 | Human | H3N2 | Throat swab | 26.7 | EX1 | OT1-MBTR, OT2-UNI, OT3-MBTA | yes |
| HU-28 | Human | H1N1pdm09 | Unknown | 27.8 | EX1 | OT1-MBTR, OT2-UNI, OT3-MBTA | yes |
| HU-29 | Human | H3N2 | Throat swab | 24.3 | EX1 | OT1-MBTR, OT2-UNI, OT3-MBTA | yes |
| HU-30 | Human | H3N2 | Throat swab | 25.0 | EX1 | OT1-MBTR, OT2-UNI, OT3-MBTA | yes |
| HU-31 | Human | H3N2 | Cell isolate | 27.6/29.2 | EX1, EX2 | OT1, OT2; MBT, UNI, MBT2 | yes |
| HU-32 | Human | H3N2 | Cell isolate | 25.5/26.5 | EX1, EX2 | OT1, OT2; MBT, UNI, MBT2 | yes |
| HU-33 | Human | H3N2 | Cell isolate | 25.3/26.6 | EX1, EX2 | OT1, OT2; MBT, UNI, MBT2 | yes |
| HU-34 | Human | H3N2 | Cell isolate | 25.5/26.7 | EX1, EX2 | OT1, OT2; MBT, UNI, MBT2 | yes |

| | | | | | | | |
|---|---|---|---|---|---|---|---|
| HU-35 | Human | H3N2 | Cell isolate | 23.4/25.2 | EX1, EX2 | OT1, OT2; MBT, UNI, MBT2 | yes |
| HU-36 | Human | H1N1pdm09 | Cell isolate | 21.3/21.9 | EX1, EX2 | OT1, OT2; MBT, UNI, MBT2 | yes |
| HU-37 | Human | H1N1pdm09 | Cell isolate | 24.2/24.2 | EX1, EX2 | OT1, OT2; MBT, UNI, MBT2 | yes |
| HU-38 | Human | H1N1pdm09 | Cell isolate | 19.5/21.9 | EX1, EX2 | OT1, OT2; MBT, UNI, MBT2 | yes |
| HU-39 | Human | H1N1pdm09 | Cell isolate | 23.4/25.6 | EX1, EX2 | OT1, OT2; MBT, UNI, MBT2 | yes |
| HU-40 | Human | H1N1pdm09 | Cell isolate | 23.6/24.1 | EX1, EX2 | OT1, OT2; MBT, UNI, MBT2 | yes |
| SW-1 | Swine | H1N1pdm09 | Pool of nasal swabs (x5) | 24.0/24.5 | EX1, EX2 | OT1, OT2; MBT, UNI | yes |
| SW-2 | Swine | H1avN2sw | Pool of nasal swabs (x5) | 23.5/23.4 | EX1, EX2 | OT1, OT2; MBT, UNI | yes |
| SW-3 | Swine | H1avN2sw | Pool of nasal swabs (x5) | 24.5/24.0 | EX1, EX2 | OT1, OT2; MBT, UNI | yes |
| SW-4 | Swine | H1N2 | Nasal swab | 28.5 | EX2 | OT1, OT2; MBT, UNI | yes |
| SW-5 | Swine | H1pdmN1pdm | Pool of nasal swabs (x4) | 30.1 | EX2 | OT1, OT2; MBT, UNI | yes |
| SW-6 | Swine | H1avN2sw | Pool of nasal swabs (x5) | 28.3 | EX2 | OT1, OT2; MBT, UNI | yes |
| SW-7 | Swine | H1N1av | Pool of nasal swabs (x5) | 26.6 | EX2 | OT1, OT2; MBT, UNI | yes |
| SW-8 | Swine | H1avN2sw | Nasal swab | 26.5 | EX2 | OT1, OT2; MBT, UNI | yes |
| SW-9 | Swine | H1pdmN1av | Nasal swab | 37.4 | EX2 | OT1, OT2; MBT, UNI | no |
| SW-10 | Swine | H1avN2sw | Pool of nasal swabs (x5) | 25.5 | EX2 | OT1, OT2; MBT, UNI | yes |
| SW-11 | Swine | H1N1 | Pool of nasal swabs (x4) | 29.1 | EX2 | OT1, OT2; MBT, UNI | yes |
| SW-12 | Swine | H1N2sw | Lung tissue | 36.2 | EX2 | OT1, OT2; MBT, UNI | no |
| SW-13 | Swine | H1avN2sw | Lung tissue | 37.1 | EX2 | OT1, OT2; MBT, UNI | no |
| SW-14 | Swine | H1N1av | Unknown | 14.6 | EX1 | OT1-MBTR, OT2-UNI, OT3-MBTA, OT4-UNI | no |
| SW-15 | Swine | H1avN2sw | Pool of nasal swabs (x5) | 26.8/25.6 | EX1, EX2 | OT1, OT2, OT5; MBT, UNI | no |
| SW-16 | Swine | H1pdmN1av | Pool of nasal swabs (x5) | 28.9/26.6 | EX1, EX2 | OT1, OT2, OT5; MBT, UNI | no |
| SW-17 | Swine | H1avN1av | Pool of nasal swabs (x5) | 26.6/25.6 | EX1, EX2 | OT1, OT2, OT5; MBT, UNI | no |
| SW-18 | Swine | H1avH1pdmN1avN2sw | Pool of nasal swabs (x5) | 26.1/25.9 | EX1, EX2 | OT1, OT2; MBT, UNI, MBT2 | yes |
| SW-19 | Swine | H1pdmN1 | Nasal swab | 34.9/36.5 | EX1, EX2 | OT1, OT2; MBT, UNI, MBT2 | yes |
| SW-20 | Swine | H3huN2 | Nasal swab | 32.0/33.1 | EX1, EX2 | OT1, OT2; MBT, UNI, MBT2 | yes |
| SW-21 | Swine | H1N2sw | Nasal swab | 27.3/27.3 | EX1, EX2 | OT1, OT2; MBT, UNI, MBT2 | yes |
| SW-22 | Swine | H1pdmN1 | Pool of nasal swabs (x5) | 38.2/35.4 | EX1, EX2 | OT1, OT2; MBT, UNI, MBT2 | yes |
| SW-23 | Swine | H1avN2sw | Pool of nasal swabs (x4) | 30.0/30.2 | EX1, EX2 | OT1, OT2; MBT, UNI, MBT2 | yes |

| | | | | | | | |
|---|---|---|---|---|---|---|---|
| SW-24 | Swine | H1N2sw | Pool of nasal swabs (x5) | 34.1/34.5 | EX1, EX2 | OT1, OT2; MBT, UNI, MBT2 | yes |
| SW-25 | Swine | H1N1 | Pool of nasal swabs (x5) | 27.0/26.0 | EX1, EX2 | OT1, OT2; MBT, UNI, MBT2 | yes |
| SW-26 | Swine | H1N2 | Cell isolate | 23.3/25.2 | EX1, EX2 | OT1, OT2; MBT, UNI, MBT2 | yes |
| SW-27 | Swine | H1N1 | Cell isolate | 24.5/26.4 | EX1, EX2 | OT1, OT2; MBT, UNI, MBT2 | yes |
| AV-1 | Avian | H5N1 | Pool of tracheal and cloacal swab | 24.9/25.5 | EX1, EX2 | OT1, OT2; MBT, UNI | yes |
| AV-2 | Avian | H9Nx | Egg culture | 23.3/24.7 | EX1, EX2 | OT1, OT2; MBT, UNI | yes |
| AV-3 | Avian | H5N1 | Brain swab | 18.7/17.8 | EX1, EX2 | OT1, OT2; MBT, UNI | yes |
| AV-4 | Avian | H9N2 | Pool of cloacal swabs | 38.5 | EX2 | OT1, OT2; MBT, UNI | no |
| AV-5 | Avian | H9N2 | Pool of cloacal swabs | 39.1 | EX2 | OT1, OT2; MBT, UNI | no |
| AV-6 | Avian | H4N6 | Pool of cloacal swabs | 36.4 | EX2 | OT1, OT2; MBT, UNI | no |
| AV-7 | Avian | H9N2 | Pool of cloacal swabs | 31.0 | EX2 | OT1, OT2; MBT, UNI | yes |
| AV-8 | Avian | H9N2 | Pool of cloacal swabs | 35.2 | EX2 | OT1, OT2; MBT, UNI | yes |
| AV-9 | Avian | H12Nx | Pool of cloacal swabs | 39.1 | EX2 | OT1, OT2; MBT, UNI | no |
| AV-10 | Avian | H9N2 | Pool of cloacal swabs | 36.6 | EX2 | OT1, OT2; MBT, UNI | no |
| AV-11 | Avian | H9N2 | Pool of cloacal swabs | 35.2 | EX2 | OT1, OT2; MBT, UNI | yes |
| AV-12 | Avian | H6N2 | Pool of cloacal swabs | 36.8 | EX2 | OT1, OT2; MBT, UNI | no |
| AV-13 | Avian | H12Nx | Pool of cloacal swabs | 38.9 | EX2 | OT1, OT2; MBT, UNI | no |
| AV-14 | Avian | H5N2 | Unknown | 17.3 | EX1 | OT1-MBTR, OT2-UNI, OT3-MBTA, OT4-UNI | no |
| AV-15 | Avian | H5N1 | Cloacal swab | 33.3/31.7 | EX1, EX2 | OT1, OT2, OT5; MBT, UNI | no |
| AV-16 | Avian | H5N1 | Cloacal swab | 33.7/29.3 | EX1, EX2 | OT1, OT2, OT5; MBT, UNI | no |
| AV-17 | Avian | H5N1 | Brain swab | 17.9/15.7 | EX1, EX2 | OT1, OT2, OT5; MBT, UNI | no |
| AV-18 | Avian | H5N1 | Pool of tracheal and cloacal swab | 27.8/29.1 | EX1, EX2 | OT1, OT2; MBT, UNI, MBT2 | yes |
| AV-19 | Avian | H5N1 | Pool of tracheal swabs | 25.2/26.3 | EX1, EX2 | OT1, OT2; MBT, UNI, MBT2 | yes |
| AV-20 | Avian | H5N1 | Brain swab | 26.2/25.6 | EX1, EX2 | OT1, OT2; MBT, UNI, MBT2 | yes |
| AV-21 | Avian | H5N1 | Brain swab | 21.6/20.5 | EX1, EX2 | OT1, OT2; MBT, UNI, MBT2 | yes |
| AV-22 | Avian | H5N1 | Pool of tracheal and cloacal swab | 24.6/29.8 | EX1, EX2 | OT1, OT2; MBT, UNI, MBT2 | yes |
| AV-23 | Avian | H5N5 | Brain swab | 21.4/21.1 | EX1, EX2 | OT1, OT2; MBT, UNI, MBT2 | yes |

| | | | | | | | |
|---|---|---|---|---|---|---|---|
| AV-24 | Avian | H5N1 | Brain | 34.3/36.3 | EX1, EX2 | OT1, OT2; MBT, UNI, MBT2 | yes |
| AV-25 | Avian | H16N3 | Egg culture | 13.7/17.3 | EX1, EX2 | OT1, OT2; MBT, UNI, MBT2 | yes |
| AV-26 | Avian | H13N5 | Egg culture | 11.6/17.7 | EX1, EX2 | OT1, OT2; MBT, UNI, MBT2 | yes |
| AV-27 | Avian | H5N2 | Egg culture | 16.4/20.4 | EX1, EX2 | OT1, OT2; MBT, UNI, MBT2 | yes |

**Table S2:** Sequences of all the primers used in the one-tube RT-PCR reactions.

| Primer name | Primer sequence (5'-3') | Used primer sets (ratio) |
|---|---|---|
| MBTUni-12(A) | ACGCGTGATCAGCAAAAGCAGG | MBT = MBTUni-12(A) / MBTUni-12(G) / MBTUni-13 (1:1:2)<br>MBT2 = MBTUni-12(A) / MBTUni-12(G) / MBTUni-13 (0.8:1.2:2)<br>MBTA = MBTUni12(A) / MBTUni13 (1:1)<br>MBTR = MBTUni12(R) / MBTUni13 (1:1) |
| MBTUni-12(G) | ACGCGTGATCAGCGAAAGCAGG | |
| MBTUni-12(R) | ACGCGTGATCAGCRAAAGCAGG | |
| MBTUni-13 | ACGCGTGATCAGTAGAAACAAGG | |
| Uni-12/Inf-1 | GGGGGGAGCAAAAGCAGG | UNI = Uni-12/Inf-1 / Uni-12/Inf-3 / Uni-13/Inf-1 (0.8:1.2:2) |
| Uni-12/Inf-3 | GGGGGGAGCGAAAGCAGG | |
| Uni-13/Inf-1 | CGGGTTATTAGTAGAAACAAGG | |

**Table S3:** Other One Tube RT-PCR methods that were tested, but did not perform better than OT1 or OT2. See **Figure S1**.

| | **OT3** (modified from Zhou et al., 2009[1]) | **OT4** (Wütrich et al., 2019[6]) | **OT5** (Galli et al., 2022) |
|---|---|---|---|
| **RNA-input** | 3 µl | 2.5 µl | 15 µl |
| **One-Step RT-PCR Polymerase (volume)** | SuperScript™ III One-Step RT-PCR System with Platinum Taq High Fidelity DNA Polymerase (Invitrogen™) (0.5 µl) | SuperScript™ III One-Step RT-PCR System with Platinum Taq High Fidelity DNA Polymerase (Invitrogen™) (1 µl) | SuperScript™ III One-Step RT-PCR System with Platinum Taq High Fidelity DNA Polymerase (Invitrogen™) (2 µl) |
| **Reaction Volume** | 25 µl | 50 µl | 60 µl |
| **Primers (Volume)** | MBTuni-12(A) (200 nM)<br>MBTuni-13 (200 nM) | Uni-12/Inf-1 (100 nM)<br>Uni-12/Inf-3 (100 nM)<br>Uni-13/Inf-1 (200 nM) | MBTuni-12(A) (500 nM)<br>MBTuni-12(G) (500 nM)<br>MBTuni-13 (1000 nM)<br>or<br>Uni-12/Inf-1 (400 nM)<br>Uni-12/Inf-3 (600 nM)<br>Uni-13/Inf-1 (1000 nM) |
| **Other reagents** | | | |
| **Thermocycler program** | 42°C for 60 min; 94°C for 2 min<br>[94°C for 30 sec; 45°C for 30 sec; 68°C for 3 min] x 5<br>[94°C for 30 sec; 57°C for 30 sec; 68°C for 3 min] x 40<br>68°C for 10 min | 42°C for 60 min; 94°C for 2 min<br>[94°C for 30 sec; 44°C for 30 sec; 68°C for 3 min] x 5<br>[94°C for 30 sec; 52°C for 30 sec; 68°C for 3 min] x 30<br>68°C for 10 min | 42°C for 60 min; 94°C for 2 min<br>[94°C for 30 sec; 45°C for 30 sec; 68°C for 3 min] x 5<br>[94°C for 30 sec; 57°C for 30 sec; 68°C for 3 min] x 35<br>68°C for 7 min |

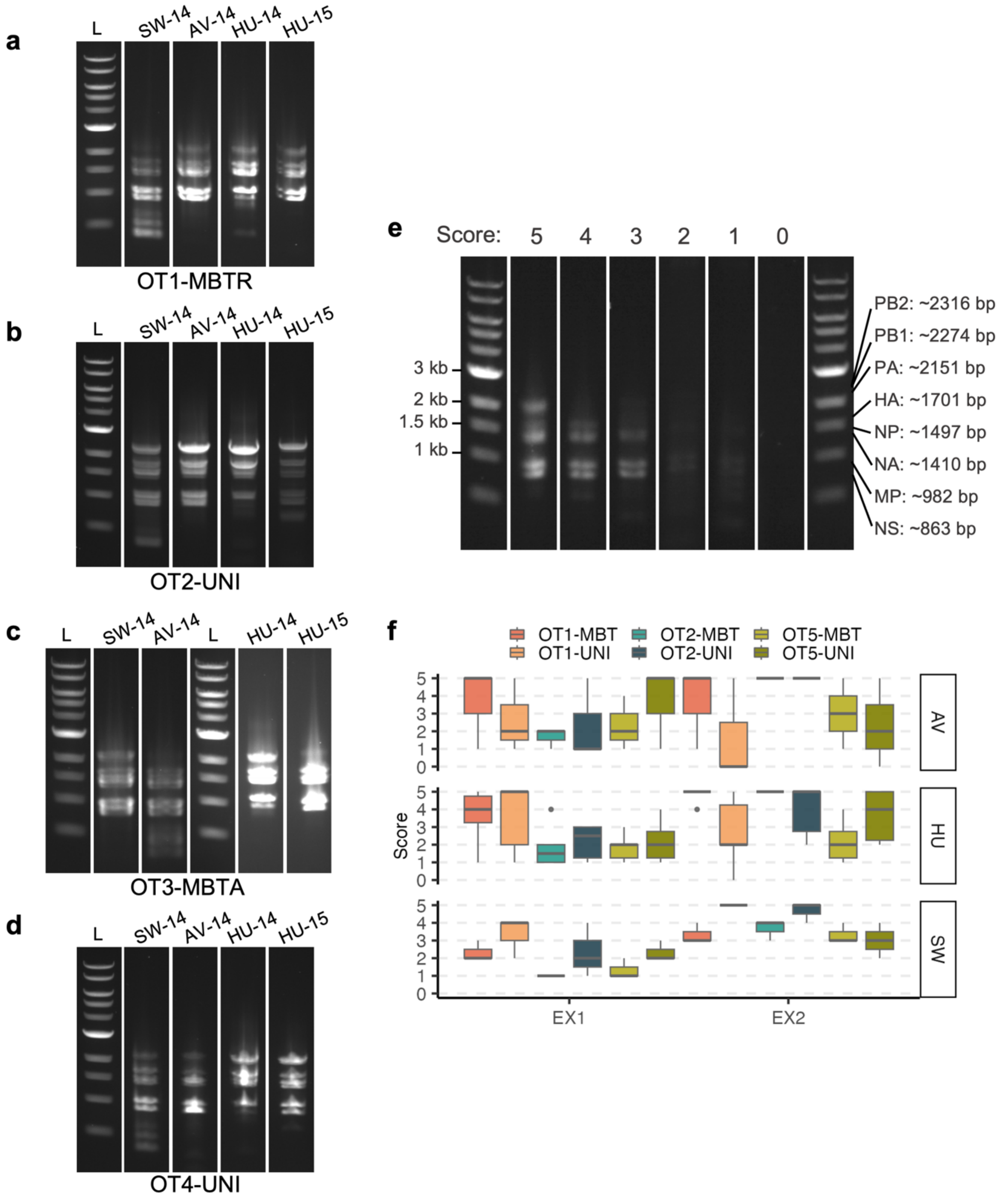


**Figure S1:** Initial comparisons of different one-tube RT-PCR protocols. **a-d:** Gel images of the PCR products of 4 different swine, avian and human origin samples amplified with **a:** OT1 with MBT12(R)/MBT13 primers; **b:** OT2 with UNI primers; **c:** OT3 with MBT12(A)/MBT13 primers; or **d:** OT4 with UNI primers. DNA ladders (L) from each gel are shown to the left of the samples. **e-f:** Initial comparisons of three one-tube RT-PCR protocols with two sets of primers using RNA from 3 avian (AV), 6 human (HU), or 3 swine (SW) origin samples extracted with either EX1 or EX2. **e.** Gel images displaying representative PCR band examples corresponding to the scorings used in **f**. The first and last lanes represent DNA ladders and have been annotated by relevant ladder fragment sizes on the left and approximate positions of PCR fragments for each IAV genome segment on the right. Scorings range from 0-5: a score of 0 indicates a blank gel, while 5 indicates clear bands for all segments (Note that the three polymerase segments, as well as NP/NA segments, are closely positioned on the gels, making them visually indistinguishable). **f.** Boxplot of the scorings of PCR products treated with different extraction and one-tube RT-PCR methods. The methods were each tested on 3 samples of avian origin, 6 samples of human origin, and 3 samples of swine origin.

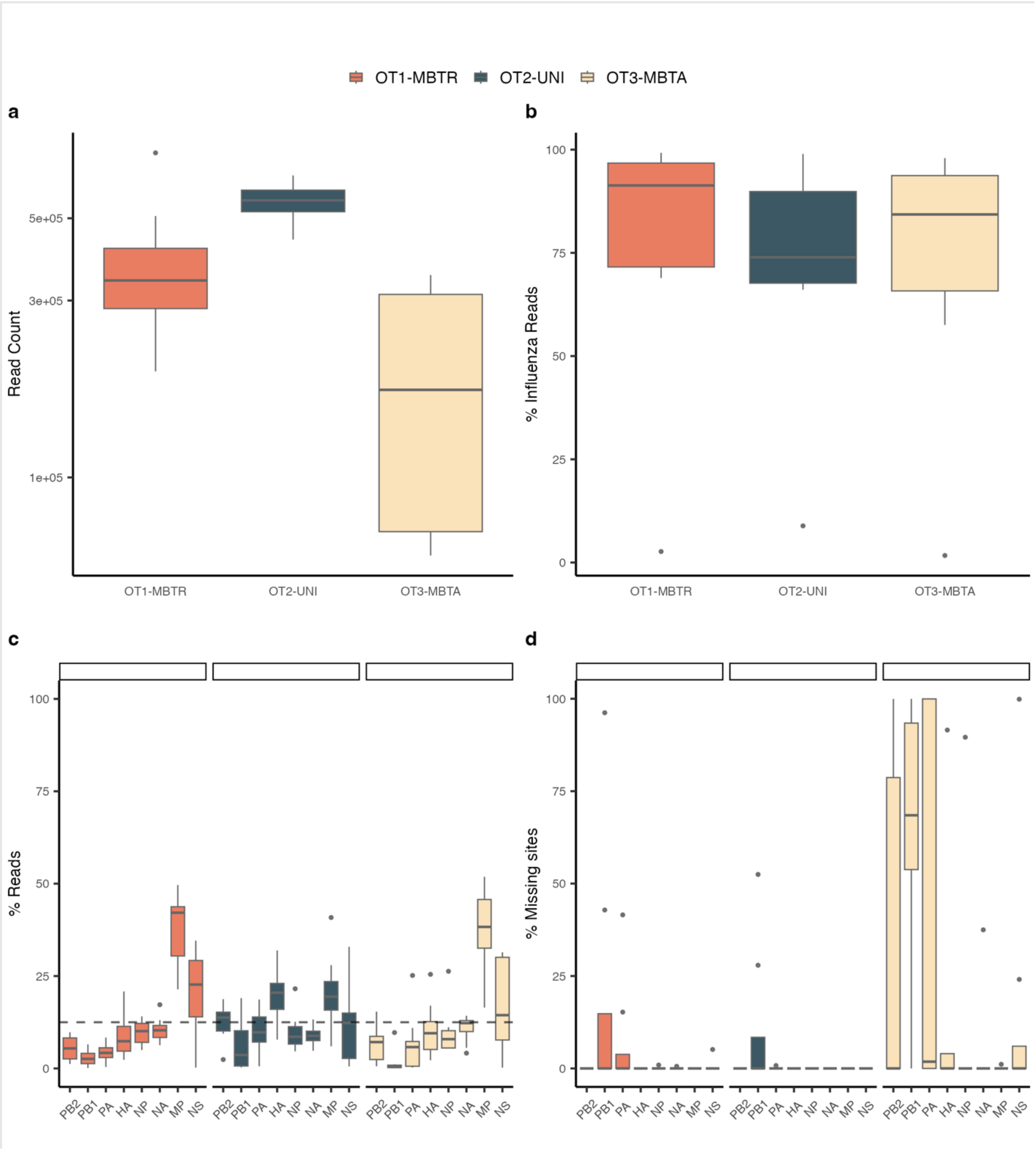


**Figure S2**: Comparisons of sequencing reads quality and distribution following either OT1 with MBT12(R)/MBT13 primers, OT2 with UNI primers or OT3 with MBT12(A)/MBT13 primers on 8 human IAV samples. **a:** Boxplot of total read counts. The y-axis is presented on a log10 scale for better visibility. **b:** Boxplot of the percentage of Influenza-specific reads. **c:** Boxplot of the distribution of reads across the viral segments calculated as the percentage of Influenza-specific reads that matched a specific segment. The dotted line at 12.5 % represents the percentage at which the distribution is equal between all eight segments. **d:** Boxplot of the percentage of missing sites (N's) assigned to the consensus sequences of each viral segment.

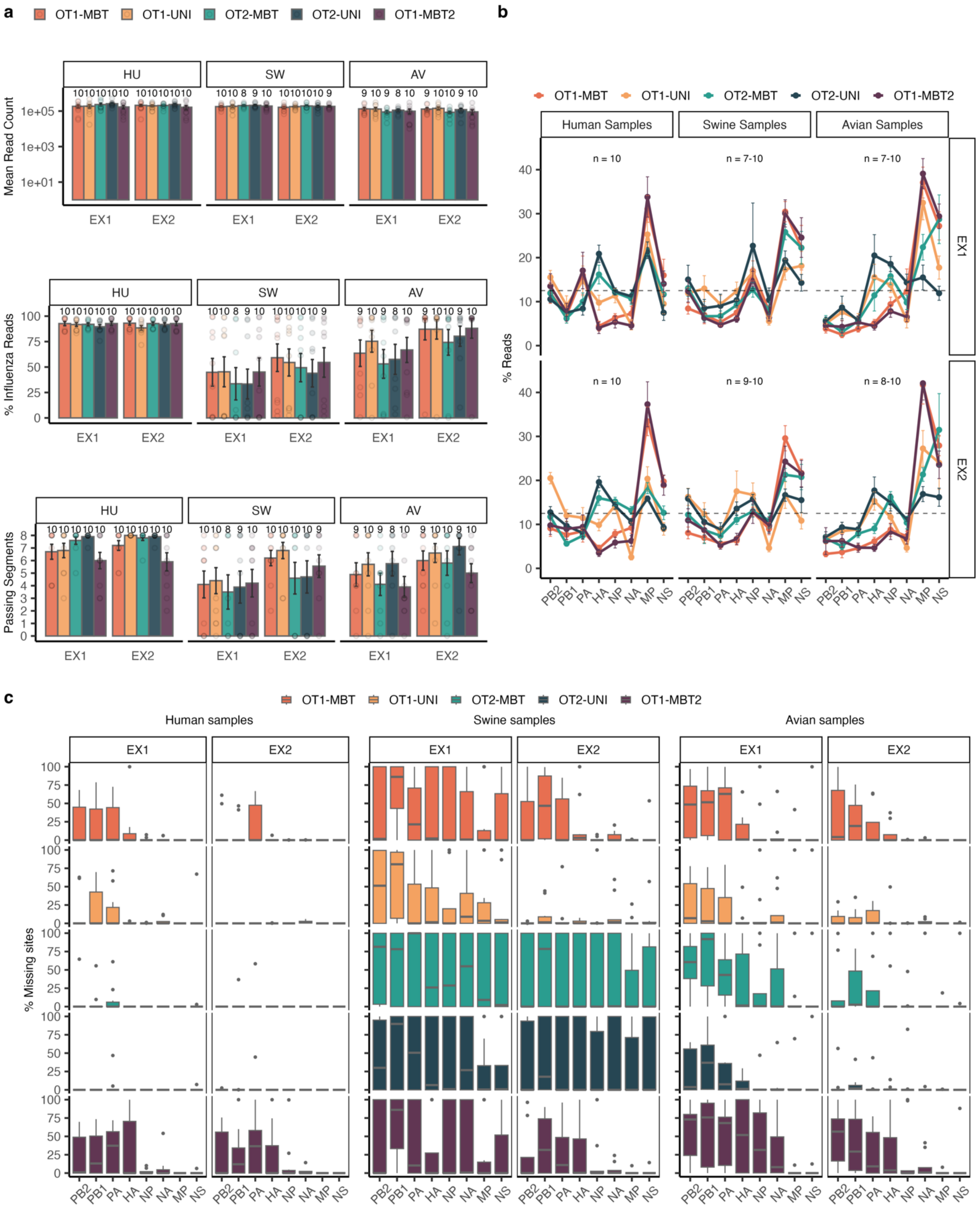


**Figure S3:** NGS read performance, distribution and quality of IAV samples of human (HU), swine (SW), or avian (AV) origin extracted using EX1 vs EX2 and amplified using two different one-tube RT-PCR methods and three different sets of primers. **a.** Boxplots of total read counts, the percentage of Influenza-specific reads, and the number of IAV segments with complete, high quality consensus sequences passing quality filters (Average read quality >30 and <10% missing bases) across all samples for each extraction method. Points represent each sample, and the total number of samples used to calculate the mean values is indicated above each bar. Error bars represent the standard error of the mean. **b.** Average percentage of influenza mapped reads in a sample that mapped to each IAV segment. Coloured lines connect the average read percentages per segment for each RT-PCR method for better visualization. The number of samples used to calculate the mean values are indicated above each plot, sometimes spanning more values when some segments were missing from one or more samples or methods. Error bars represent the standard error of the mean, and the dotted line indicates the 12.5% resulting from an equal distribution of reads to each of the 8 segments. **c**. Boxplot showing the percentage of missing sites in consensus sequences of each IAV segment. The number of samples included is not shown on the boxplots, but is similar to the numbers in **b**; Human samples EX1: n = 10, EX2: n = 10; Swine samples EX1: n = 7-10, EX2: n = 9-10; Avian samples EX1: n = 7-10, EX2: n = 8-10.

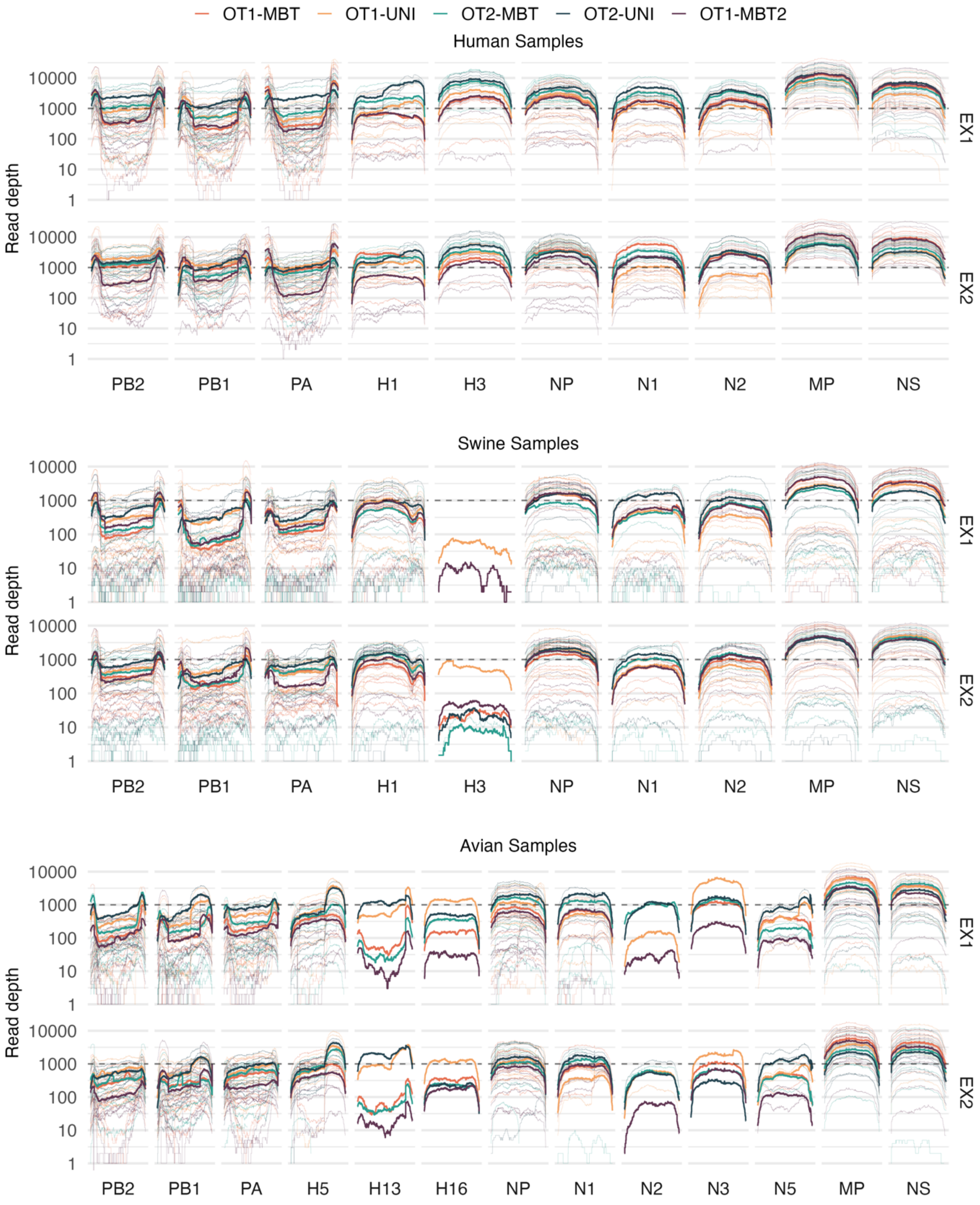


**Figure S4**: Coverage plots showing the number of reads at each position of each segment for all samples extracted using EX1 or EX2 and amplified with two different one-tube RT-PCR methods and 3 different sets of primers. Thin lines each represent the coverage of one sample for each segment, while the thick lines represent the average coverage observed for all samples treated with a specific one-tube RT-PCR method. The y-axis is presented on a log10 scale, and a dotted line shows a read depth of 1000 reads for better visibility and comparison between segments.
